# Mediating effects of inequitable gender norms on intimate partner violence and contraceptive use in a cluster randomized control trial in Niger: A causal inference mediation analysis

**DOI:** 10.1101/2023.01.12.23284504

**Authors:** Sabrina C Boyce, AM Minnis, J Deardorff, SI McCoy, DE Goin, S Challa, NE Johns, S Aliou, MI Brooks, A-M Nouhou, H Baker, JG Silverman

## Abstract

**Background:** Gender inequity, a deeply-rooted driver of poor health globally, is expressed in society through gender norms, the unspoken rules that govern gender-related roles and behavior. The development of public health interventions focused on promoting equitable gender norms are gaining momentum internationally, but there remain critical gaps in the evidence about how these interventions are working to change behavioral outcomes.

**Methods:** A four-arm cluster randomized control trial (cRCT) was conducted to evaluate the effects of the Reaching Married Adolescents in Niger (RMA) intervention on modern contraceptive use and intimate partner violence (IPV) among married adolescent girls and their husbands in Dosso, Niger (T1: 1042 dyads; 24 mos. follow-up: 737 dyads, 2016-2019). This study seeks to understand if changes in perceived inequitable gender norms among husbands are the mechanism behind effects on modern contraceptive use and IPV. We estimated natural direct and indirect effects via these gender norms using inverse odds ratio weighting. An intention-to-treat approach and a difference-in-differences estimator in a hierarchical linear probability model was used to estimate prevalence differences, along with bootstrapping to estimate confidence intervals.

**Results:** The total effects of the RMA small group intervention (Arm 2) is estimated to be an 8% reduction in prevalence of IPV [95% CI: -0.18, 0.01]. For this arm, the natural indirect effect through gender inequitable social norms is associated with a 2% decrease (95% CI: -0.07, 0.12), accounting for 22.3% of this total effect, and the natural direct effect with a 6% decrease (95% CI: -0.20, -0.02) in IPV. Of the total effect of the RMA household visit intervention (Arm 1) on contraceptive use (20% increase), indirect effects via inequitable gender norms were associated with an 11% decrease (95% CI: -0.18, -0.01) and direct effects with a 32% increase (95% CI: 0.13, 0.44) in contraceptive use. For the combination arm, of the total effects on contraceptive use (19% increase), indirect effects were associated with a 9% decrease (95% CI: -0.20, 0.02) and direct effects with a 28% increase (95% CI: 0.12, 0.46).

**Conclusion:** The present study contributes experimental evidence that the small group RMA intervention reduced IPV partially via reductions in perceived inequitable gender norms among husbands. Evidence also suggests that increases in perceived inequitable gender norms resulted in decreased contraceptive use among those receiving the household visit intervention component. Not only do these results open the “black box” around how the RMA small group intervention may create behavior change to help inform its future use, they provide evidence supporting behavior change theories and frameworks that postulate the importance of changing underlying social norms in order to reduce IPV and increase modern contraceptive use.

## Introduction

Gender inequity is a deeply-rooted driver of poor health, including violence against women and poor reproductive health, globally (1). This pattern is clear in countries like Niger, which ranks 154^th^ of 162 nations on the United Nations (UN) Gender Inequality Index and has some of the highest rates of maternal mortality (509 per 100,000 live births), fertility (6.9 births per woman), and girl child marriage (76% of girls) in the world (2-5). The United Nation’s Sustainable Development Goals, as well as many of the most influential international development leaders, have included gender equity and empowerment of women and girls as critical to improving global health, well-being, and development (6, 7). Gender inequity is considered to be a social determinant of health that, if changed, could have long-term, expansive impact across a wide range of health outcomes (1). The development of public health interventions addressing structural and social determinants of gender inequity are gaining momentum internationally, but there remain critical gaps in the evidence about what works to change gender inequity (8).

Social norms are a critical pathway through which gender inequity constrains health (1). Gender inequity is often expressed through gender norms, a subset of social norms that constitute the unspoken and unwritten rules that govern gender-related roles and behavior (9). Violation of these rules triggers social sanctions, while compliance triggers rewards to motivate adherence (9). Understood through social norms theory, the theory of gender and power, and feminist theory, gender norms are upstream, culturally-bound determinants of many health behaviors, including those related to gender-based violence, HIV, reproductive health, and female genital cutting, to name only a few (10-13). Various conceptual frameworks for addressing health outcomes related to gender inequity position gender norms as central to improving health (8, 13, 14). Correlational evidence suggests a strong relationship between social norms and various gender inequity-related health outcomes, but does not causally demonstrate that inequity-related health outcomes are reduced by shifting toward more gender equitable norms (15-17).

Randomized evaluations have the potential to produce causal evidence to understand if social norms-focused public health interventions successfully change entrenched norms and if changes in these norms are one mechanism through which health outcomes are achieved. Understanding the mechanism of change behind interventions aiming to improve the entrenched health effects of gender inequity is critical for unpacking how interventions influence outcomes. Moreover, clarifying the mechanisms of change may shed light on whether they are temporarily shifting behavior for a short period via more proximal determinants (such as attitudinal change) or changing norms themselves, an underlying, upstream determinant of health. If the latter, the benefits could be long-lasting and include a wide range of health outcomes, possibly meriting prioritization of such interventions in resource-limited contexts, such as low- and middle-income countries (LMIC) (1, 18). Very few program evaluations conducted in LMIC contexts, however, provide evidence of the direct link between programming and normative change as determinant of health behavior change (10, 19, 20). While interventions seeking to shift social norms, often via community mobilization, small group or community discussions, and/or social norms campaigns, have demonstrated positive effects on either social norms or gender-inequity related health outcomes, very few have assessed the mechanism through which these health changes have occurred (21-27). No identified randomized evaluations of gender norm-focused interventions in LMIC have both assessed for mechanistic effects and provided evidence of health improvements occurring via change in the targeted social norms.

This study provides an opportunity to understand whether change in gender norms is a mechanism through which the Reaching Married Adolescents in Niger (RMA) intervention reduced intimate partner violence (IPV) and increased modern contraceptive use among married adolescents and their husbands. The RMA intervention included two main components – household visits and small group discussions - and was evaluated using a 4-arm factorial cluster randomized control trial in the Dosso region of Niger. Previous work has demonstrated evidence of an effect of the RMA household visit intervention on inequitable gender norms and modern contraceptive use and of the RMA small group intervention on inequitable gender norms and IPV (28, 29). This study aims to examine changes in inequitable gender norms as a pathway through which household visits increased modern contraceptive use, as well as through which small group discussions reduced IPV, using a causal inference mediation approach.

## Methods

### Intervention

The RMA intervention is a gender-synchronized (i.e., concurrently delivered to husbands and wives), community health worker intervention developed by Pathfinder International in Dosso, Niger from 2017-2019. Male and female community health workers (CHW) were trained to facilitate individual-level household visits (delivered monthly to husbands and wives separately) and single-sex small group discussions (delivered twice per month for wives and monthly for husbands) that provided information about and access to a range of family planning methods and focused on the benefits of healthy birth spacing (i.e., at least two years between births)(30). Content delivered by CHWs included: general health and life skills; reproduction anatomy and health information, including on modern contraceptive methods as a way to achieve healthy timing and spacing of pregnancies; discussion of gender norms that impede contraceptive use and female autonomy, couples’ communication regarding fertility decisions, and the harmful effects of gender-based violence. Additionally, village-level community dialogues with leaders and gatekeepers were implemented to create a supportive community environment around healthy birth spacing, including modern contraceptive use among married adolescent girls and their husbands. Details on the intervention and research protocols have been published previously (31).

### Study Design & Sample

#### Study Design

A four-arm cluster randomized control trial (cRCT) was conducted to evaluate the effects of the individual and combined intervention components. The four arms included 1) individual household visits (Arm 1), 2) small group discussions (Arm 2), 3) both household visits and small group discussions (combination; Arm 3), and 4) control (Arm 4), with all intervention arms also receiving community dialogues. Contraceptive products were stocked at local health centers in all study arms. The study was conducted in a rural region of Niger called Dosso. Three health districts within Dosso were selected by the Ministry of Health for inclusion in the study: Dosso, Doutchi, and Loga. Each district was randomly assigned one of three treatment conditions (stage 1 of randomization; 1:1:1). Within each district, 16 eligible villages [having at least 1000 residents; primarily Hausa or Zarma-speaking (the two major languages of central southern Niger); located within 5km of a health center; and having received no intervention specific to contraceptive use or gender equity] were randomly selected for inclusion in the study; 12 were randomly assigned to receive the designated treatment condition for that district and 4 to control based on computer-generated random selection of sequential numbers assigned to villages (stage 2 of randomization; 1:1:1:1). Within each village, village leaders provided a registry of all households including a married female adolescent age 13-19 years old and 25 of these households were randomly selected for recruitment using a similar computer-generated random number selection process. Additional participant eligibility included: 1) being Hausa or Zarma speaking; 2) not planning to move away from the village in next 18 months; and 3) not planning to travel away from the village for more than 3 months during that period; 4) not sterilized (though sterilization is illegal in this setting); and 5) providing informed consent. Those households that did not meet these additional criteria or were unavailable after three attempts were replaced by another household randomly selected from the registry. Baseline data were collected May through July 2016 (T1) and the same households were revisited at 24 months (April through June 2018; T2), after 12 months of intervention implementation (April 2017-February 2018). Sex-matched trained research assistants who spoke Hausa and/or Zarma recruited participants at their household and conducted the interviewer-administered 40-60 minute survey with husbands and wives separately, in locations determined by the participants and research assistant to provide auditory privacy. Responses were entered into tablet computers where data were securely uploaded at regular intervals to a secure network.

This trial was pre-registered on clinicaltrials.gov (NCT03226730), with contraceptive use defined as the primary outcome and IPV and social norms regarding gender equity defined as secondary outcomes. Study protocols were approved by the ethics review boards of the University of California San Diego School of Medicine and the Niger Ministry of Health.

#### Sample

At T1, 1351 eligible husband-wife dyads were invited to participate in the research. Surveys were collected from 1072 adolescent wives (79.3% female participation), 968 of whom also provided survey data at T2 (90.3% female retention), and from 1080 husbands (79.9% male participation), of whom 773 also participated in data collection at T2 (71.6% male retention). In Arm 1, 50.3% of husbands participated in at least one program element, 47.0% in Arm 2, and 48.7% in Arm 3 (29).

We previously reported baseline imbalances across study arms (29). Arm 1 had lower levels of Quranic education (i.e., theological education on the Qur’an), total household assets, perceived inequitable gender norms by husbands relative to controls (score of 3.7 out of 5 for Arm 1 vs. 4.3 for control arm; p<0.001, *data not shown*). Arm 2 had higher levels of government and Quranic education, lower levels of husband migration; husbands were older and wives younger at baseline, and wives were younger when married relative to controls. Arm 3 had lower levels of Quranic education and total household assets relative to controls.

### Measurements

#### Outcomes

Outcomes were measured at T2. Current modern contraceptive use was reported by wives and measured as an affirmative response to currently using an intrauterine device, injectable, implant, contraceptive pill, male condom, female condom, emergency contraception, and/or lactational amenorrhea (LAM) to delay or limit their number of pregnancies. Women who were currently pregnant were excluded from models using this outcome; we did not exclude women who reported they were trying to get pregnant because strong social expectations of fertility among young, nulliparous women in Niger may obscure any distinction among these adolescent wives and their willingness to use contraception (32). IPV, physical and/or sexual violence from husbands, in the past 12 months was also reported by wives and measured using eight items from the Demographic Health Survey (DHS) domestic violence module (33). Physical violence items included a) pushed her, shaken her or thrown something at her; b) slapped her; c) twisted her arm or pulled her hair; d) hit her with his fist or something that could hurt her; e) kicked her, dragged her, or beat her up; f) choked her or tried to burn her. Adolescent wives were also asked whether their husband had physically forced them to a) have sexual intercourse when she did not want to, or b) to perform any other sexual acts she did not want to. An affirmative response by wives to one or more items was dichotomously categorized as having experienced past 12-month IPV. Study protocols followed the World Health Organization’s guidelines for conducting research on violence against women (e.g., only one woman or girl per household was asked these questions, men were not asked these questions, and audio privacy was provided) to protect the safety and confidentiality of women and girls participating in the study (34).

#### Mediator

Social norms regarding gender equity were measured with husbands at T2 using an adaptation of the Gender Equitable Men Scale (GEMS), a validated scale to assess individual attitudes about gender that has been used in over 20 countries (35). The question stem, “People in your community think that…” was added to five items from the GEM scale to assess second order beliefs social norms. The items then proceeded as, 1) “…a woman’s most important role is to take care of the home and cook for the family”; 2) “…a man should have the final word about decisions in the home”; 3) “…there are times when a woman deserves to be beaten”; 4) “…a woman should never question her husband’s decisions even if she disagrees with them”; 5) “…it is natural and right that men have more power than women in the family.” Response options were coded dichotomously as 1 if “agree” or “somewhat agree” and 0 if “disagree”, and missing for “don’t know”/ “decline to answer”/missing responses. Response values were summed across all 5 items to create a score (Cronbach’s alpha = 0.62, range: 0-5). A total score of five indicated some or full agreement with all 5 items. Scores were dichotomized as having a score of 5 (high inequitable gender norms) vs. less than 5 (lower inequitable gender norms) in order to provide insight on shifts in high levels of social acceptability of gender inequity. Social norms were measured contemporaneously with outcomes. Because no data were collected between T1 and T2, temporality was preserved between the exposure and the mediator, but not between the mediator and outcomes.

#### Covariates

Covariates were included in all analytic models to control for factors related to baseline imbalances across study arm that could create confounding associations and factors related to couple loss-to-follow-up to reduce selection bias. In the total and natural direct and indirect effects models, covariates were included that were associated with baseline socio-demographic imbalances across the study arms (assessed via univariate multinomial regression that accounted for clustering, p<0.05) and hypothesized, based on content expertise and theory, to also relate to the outcomes. These covariates included district, wife age, husband and wife education (modern, Quranic, or no education), husband migration for three or more months in the previous year (yes, no), and total household assets. Total household assets were assessed as a count of six household items, a measure of wealth used by the Niger DHS (33). In the model used to develop mediation weights, we included covariates that were associated with baseline imbalances across the study arm (p<0.05) and hypothesized, based on theory and content expertise, to also relate to the mediator, which was husband perceived inequitable gender norms at T2. These included husband and wife education, husband migration, and total household assets (29). In the model used to develop inverse probability of censoring weights to account for loss-to-follow-up, covariates were included that were associated with loss-to-follow-up for husband-wife dyadic pairs (assessed via t-tests for continuous variables and fisher’s exact for categorical variables, p<0.10): husband current age, husband and wife ages at marriage, husband and wife parity, wife education, and husband migration.

### Statistical Analysis

#### Main Effects

Previously reported results provide evidence that receipt of the RMA household visits (Arm 1) and combination (Arm 3) were associated with an increase in current modern contraceptive use, and receipt of the RMA small group visit (Arm 2) was associated with a decrease in IPV (28). Therefore, in this analysis, only results of the mediation analyses for Arm 1 and 3 will be reported for the contraceptive use outcome and those of Arm 2 for the IPV outcome (Figure 1). Total effects for each outcome were assessed using an intention-to-treat approach and a difference-in-differences estimator to further account for baseline imbalances across study arms. A mixed-effects linear probability model was utilized to estimate prevalence differences with clustering at the village level and to account for multiple observations on each couple. Inverse probability of censoring weights (IPCW) were included to reduce selection bias related to couple loss-to-follow-up (36). For this analysis, we utilized linear models to estimate prevalence differences to accommodate the assumptions of the difference in differences estimator, rather than the incidence rate ratios estimated in the original report (28).

**Figure 1.**
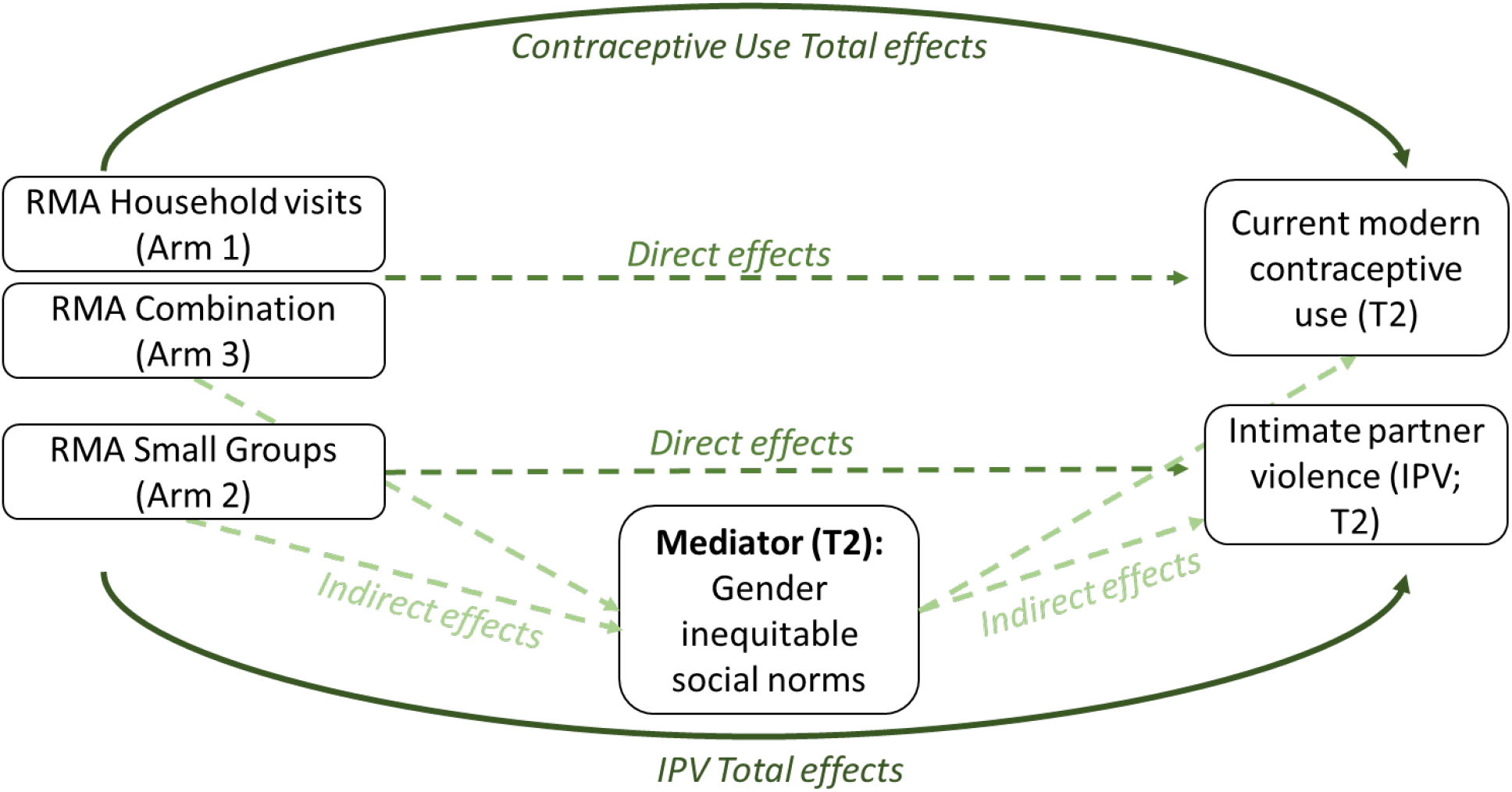
Hypothesized mediation pathway between the Reaching Married Adolescents in Niger (RMA) Intervention, gender inequitable social norms, and two outcomes, intimate partner violence (IPV) and modern contraceptive use (Dosso, Niger; 2016-2019)

#### Mediation

Previously, we reported that assignment to the RMA small group intervention was associated with a reduction in perceived inequitable gender norms, the household visits with a modest increase in perceived inequitable gender norms, and no effect for the combination arm (29). To assess the next stage of the hypothesized mediation pathway, we assessed the relationship between inequitable gender norms and both primary outcomes, contraceptive use and IPV, using a linear mixed effects model to estimate prevalence differences.

To estimate natural direct and indirect effects via these gender norms, we utilized inverse odds ratio weighting (37, 38). First, we estimated the natural direct effect, which is the pathway between the RMA intervention and outcome that does not include the mediator. Using the inverse odds ratio weighting approach, which utilizes the invariance property of the odds ratio, we estimated the odds of being included in each study arm based on the mediator and potential confounders of the exposure-mediator relationship using a multinomial regression model that included the IPCW weights (39). The inverse of the predicted probabilities from this model were then used to create the stabilized mediation weights, which were included in the total effects model to estimate the natural direct effect by blocking the indirect pathway between the RMA intervention and gender norms.

Natural indirect effects estimate the hypothesized pathway between the RMA intervention and outcome that travels through inequitable gender norms (Figure 1). Because the natural direct and indirect effect estimates sum to equal the total effects, the natural indirect effect is calculated by subtracting the natural direct effect estimate from that of the total effect. The nonparametric bootstrap accounting for village-level clustering was used to calculate 95% Wald-type confidence intervals for all effect estimates (n=1,000). All analyses were conducted using RStudio, version 2022.02.1 build 461 (40).

## Results

Sociodemographic characteristics of the husband-wife dyads, overall and by the two outcomes, are presented in Table 1. For this analysis, 1042 husband-wife dyads were included from T1 and 737 husband-wife dyads from T2. At baseline, husbands were 25.6 years and wives were 17.3 years on average. Husbands reported having 1.5 children, while adolescent wives reported 1.0 (the difference related to polygamous marriages, 13% of husbands). Among husbands, 45.2% perceived high levels of gender inequitable social norms at T1 and 37.9% at T2. In the overall sample, 8.7% of married adolescent girls reported experiencing IPV from their husbands in the past 12 months at T1 and 9.7% at T2. Additionally, 11.8% reported currently using a form of modern contraceptive at T1 and 38.3% at T2.

**Table 1.** Baseline covariates and husband perceived social norms (T2) by past 12-month intimate partner violence and current modern contraceptive use (T2) among female married adolescent girls and their husbands in Niger (Dosso, Niger; 2016-2019)

|  | Overall<br>(n=1042) | Intimate Partner<br>Violence<br>(Past 12 months; T2)<sup>a</sup> |  | Current Modern<br>Contraceptive Use (T2)<sup>a</sup> |  |
| --- | --- | --- | --- | --- | --- |
|  |  | Yes (n=92) | No (n=825) | Yes (n=301) | No (n=496) |
| Study arm (%) |  |  |  |  |  |
| Household Visits (Arm 1) | 279 (26.8) | 23 (25.0) | 223 (27.0) | 81 (26.9) | 129 (26.0) |
| Small Groups (Arm 2) | 247 (23.7) | 16 (17.4) | 199 (24.1) | 75 (24.9) | 112 (22.6) |
| Combination (Arm 3) | 256 (24.6) | 26 (28.3) | 201 (24.4) | 85 (28.2) | 118 (23.8) |
| Control | 260 (25.0) | 27 (29.3) | 202 (24.5) | 60 (19.9) | 137 (27.6) |
| District (%) |  |  |  |  |  |
| Dosso | 342 (32.8) | 35 (38.0) | 269 (32.6) | 100 (33.2) | 170 (34.3) |
| Doutchi | 335 (32.1) | 26 (28.3) | 268 (32.5) | 96 (31.9) | 157 (31.7) |
| Loga | 365 (35.0) | 31 (33.7) | 288 (34.9) | 105 (34.9) | 169 (34.1) |
| **Baseline Covariates** | **mean (SD)** | **mean (SD)** | **mean (SD)** | **mean (SD)** | **mean (SD)** |
| Husband age | 25.6 (5.4) | 25.2 (5.0) | 25.7 (5.4) | 26.2 (5.6) | 25.4 (5.2) |
| Husband age at marriage | 22.5 (5.0) | 22.5 (4.7) | 22.4 (5.0) | 22.8 (5.3) | 22.3 (4.8) |
| Husband parity | 1.5 (2.0) | 1.4 (2.1) | 1.5 (2.0) | 1.7 (2.1) | 1.3 (2.0) |
| Husband education (%) |  |  |  |  |  |
| Any modern | 502 (48.5) | 45 (48.9) | 396 (48.4) | 170 (57.0) | 222 (45.0) |
| Quaranic only | 215 (20.8) | 20 (21.7) | 174 (21.3) | 49 (16.4) | 111 (22.5) |
| No schooling | 318 (30.7) | 27 (29.3) | 248 (30.3) | 79 (26.5) | 160 (32.5) |
| Husband migration for >3 months (%) | 722 (69.8) | 67 (73.6) | 560 (68.4) | 193 (64.8) | 356 (72.2) |
| Wife age | 17.3 (1.5) | 17.4 (1.4) | 17.3 (1.6) | 17.4 (1.5) | 17.2 (1.6) |
| Wife age at marriage | 14.2 (1.9) | 14.5 (1.9) | 14.1 (1.9) | 14.1 (1.8) | 14.2 (1.9) |
| Wife parity | 1.0 (1.0) | 0.9 (0.9) | 1.0 (1.0) | 1.1 (1.0) | 0.9 (1.0) |
| Wife education (%) |  |  |  |  |  |
| Any modern | 364 (35.1) | 46 (50.0) | 275 (33.6) | 137 (45.7) | 150 (30.4) |
| Quaranic only | 171 (16.5) | 15 (16.3) | 142 (17.3) | 36 (12.0) | 96 (19.5) |
| No schooling | 501 (48.4) | 31 (33.7) | 402 (49.1) | 127 (42.3) | 247 (50.1) |
| Total household assets | 2.1 (1.2) | 2.0 (1.1) | 2.1 (1.2) | 2.0 (1.2) | 2.1 (1.2) |
| **Mediator** | **n (%)** | **n (%)** | **n (%)** | **n (%)** | **n (%)** |
| High perceived inequitable gender norms |  |  |  |  |  |
| T1 (n=1042) | 460 (45.2) | 48 (53.3) | 359 (44.5) | 127 (42.8) | 229 (47.6) |
| T2 (n=737) | 275 (37.9) | 36 (46.8) | 231 (37.4) | 81 (32.4) | 151 (42.3) |

Abbreviations: T1, baseline; T2, 24-month follow-up; SD, standard deviation
<sup>a</sup>Intimate partner violence and modern contraceptive use based on wife reports at T2

### Baseline imbalances of outcomes across study arms

At baseline, wives in Arm 3 reported significantly higher levels of IPV relative to those in the control arm (14.8% vs. 6.9%, p=0.01); no significant differences were observed for other study arms. Also at baseline, wives in Arm 1 and 3 reported significantly lower levels of current modern contraceptive use relative to control (Arm 1: 5.7%, Arm 3: 7.0%, Control: 14.2%, p<0.01); no significant difference was observed for Arm 2. Other imbalances in sociodemographic characteristics across study arms are reported elsewhere (28, 29).

### Baseline characteristics associated with couple retention at T2

Several sociodemographic differences were observed between couples retained at T2 and those lost-to-follow-up, however, the magnitude of these differences was modest and likely not practically relevant. Couples retained in the study at T2 were more likely to have husbands who were older at the time of the study (25.9 years among those retained vs. 25.0 years among those not retained, p=0.01) and older when married (22.6 year vs. 22.1 years, p=0.09), with wives who were younger when married (14.1 years vs. 14.3 years, p=0.05), relative to those couples not retained at T2 (data not shown). Retained couples also had significantly more children at baseline compared to non-retained couples, with wives from retained couples reporting a mean of 0.99 children (vs. 0.87 among non-retained, p=0.07) and with husbands reporting a mean of 1.60 children (vs. 1.26 among non-retained, p=0.01).

### Main effects

Estimates of the total effects of the RMA intervention indicate a null effect of the household visit intervention (Arm 1) on IPV prevalence [adjusted prevalence difference (aPD): 0.01, 95% CI: -0.08, 0.08), that assignment to the small group intervention (Arm 2) is associated with an 8% reduction in prevalence of IPV (aPD: -0.08, 95% CI: -0.18, 0.01) (Table 2), and assignment to the combination arm (Arm 3) with a 10% reduction in IPV (aPD: -0.10, 95% CI: -0.18, 0.02). For modern contraceptive use, total effect of the household visits (Arm 1) is a 20% increase in use (aPD 0.20, 95% CI: 0.03-0.36), for the small group intervention (Arm 2) an 11% increase (aPD: -0.01, 0.24), and for the combination (Arm 3) an 19% increase in current modern contraceptive use (aPD 0.19, 95% CI: 0.07, 0.34) (Table 2).

**Table 2.** Estimated total, natural direct and indirect effects^a^ of the Reaching Married Adolescents in Niger intervention on past 12-months intimate partner violence and current modern contractive use via perceived inequitable gender norms among husbands of married adolescents (Dosso, Niger; 2016-2019)

|  | Total Effect |  | Natural Direct Effect |  | Natural Indirect Effect |  |
| --- | --- | --- | --- | --- | --- | --- |
|  | aPD<sup>b</sup> | 95% CI<sup>c</sup> | aPD<sup>b</sup> | 95% CI<sup>c</sup> | aPD<sup>b</sup> | 95% CI<sup>c</sup> |
| **Intimate partner violence (past 12-months) (n=1000 couples)** |  |  |  |  |  |  |
| Arm 2: Small Groups<sup>d</sup> | -0.08 | -0.18, 0.01 | -0.06 | -0.20, -0.02 | -0.02 | -0.07, 0.12 |
| **Current modern contraceptive use (n=970 couples)** |  |  |  |  |  |  |
| Arm 1: Household Visits<sup>d</sup> | 0.20 | 0.03, 0.36 | 0.32 | 0.13, 0.44 | -0.11 | -0.18, -0.01 |
| Arm 3: Combined<sup>d</sup> | 0.19 | 0.07, 0.34 | 0.28 | 0.12, 0.46 | -0.09 | -0.20, 0.02 |

Abbreviations: aPD, adjusted prevalence difference; CI, confidence interval
<sup>a</sup>Natural direct and indirect effects were estimated using inverse odds ratio weighting (Tchetgen, Tchetgen 2013).
<sup>b</sup>aPD estimated using a mixed effects difference-in-difference linear regression with inverse probability of censoring weights and adjusting for household assets, husband education, wife education, and husband migration with clustering based on village and multiple observations
<sup>c</sup>Wald-type 95% CI estimated using nonparametric bootstrap with 1000 repetitions
<sup>d</sup>Only arms for which significant effects on main outcomes were detected were included in this table and in mediation analyses.

### Associations between inequitable gender norms and main outcomes

The effects of each intervention arm on inequitable gender norms, the first stage of the mediation pathway, have been previously reported; the small group intervention was associated with a reduction in inequitable gender norms, household visits with a moderate increase in inequitable gender norms, and the combination arm was not associated with these norms (29). The second stage of the mediation pathway is the relationship between inequitable gender norms and the main outcomes, contraceptive use and IPV. Among husbands’ who perceived high inequitable gender norms, the prevalence of IPV victimization among their wives is 5% higher (aPD: 0.05, 95% CI: -0.002, 0.09) the prevalence of modern contraceptive use among their wives is 7% lower (aPD: -0.07; 95% CI: -0.14, 0.01), than among husbands who perceived lower levels of inequitable norms (Figures 2 and 3).

**Figure 2.**
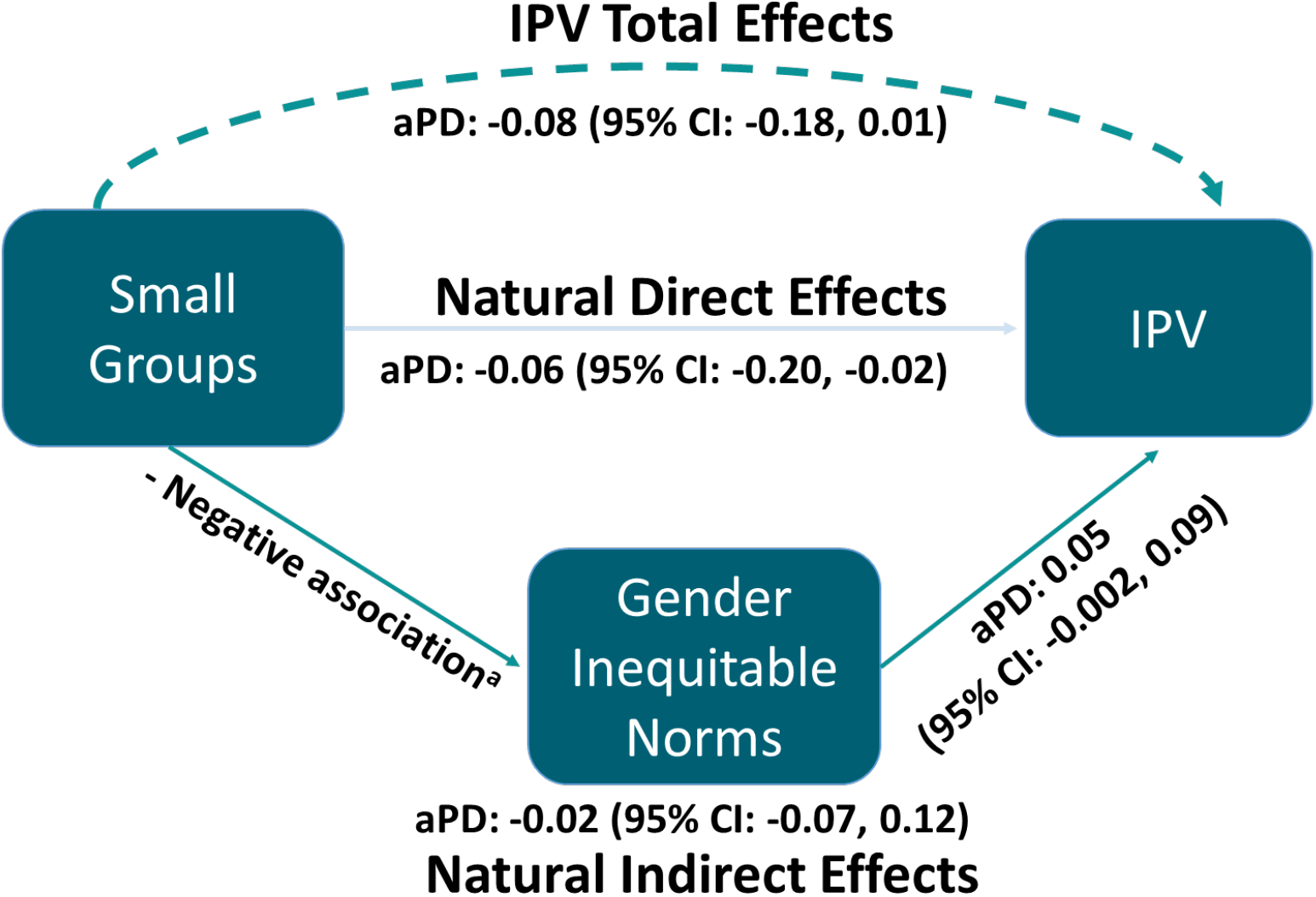
Estimated total, natural direct and indirect effects of the Reaching Married Adolescents in Niger small group intervention on intimate partner violence via perceived inequitable gender norms among husbands of married adolescent girls in Dosso, Niger at 24-month follow-up (2017-2019) ^a^**Boyce SC**, Deardorff J, Minnis AM, McCoy S, Shakya H, Challa S, Brooks M, Aliou S, NouhouA, Silverman JG. Effect of a small group gender-synchronized family planning intervention on perceived gender norms among husbands of married adolescent girls in a cluster randomized control trial in Dosso, Niger. *Under review*.

**Figure 3.**
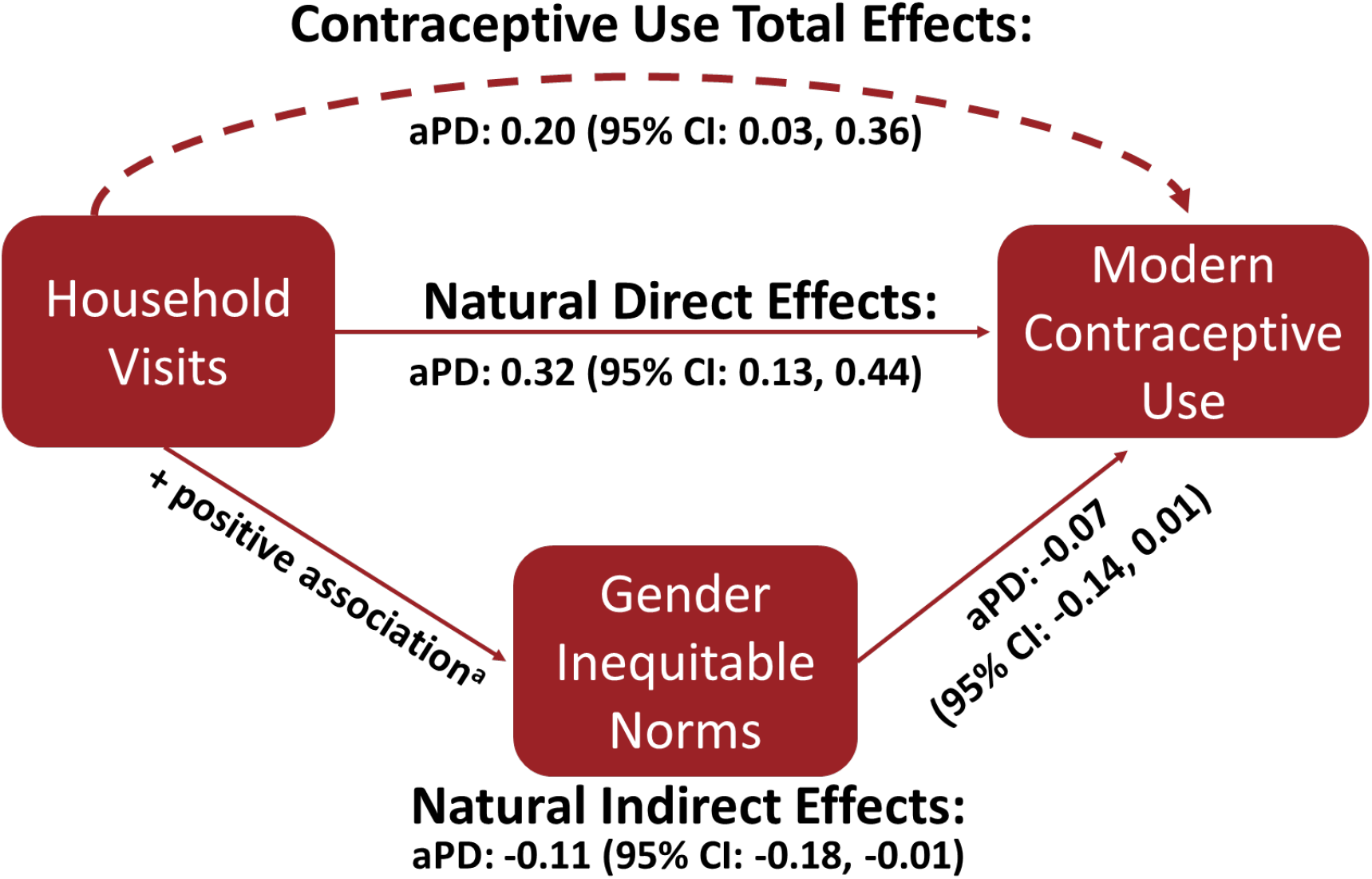
Estimated total, natural direct and indirect effects of the Reaching Married Adolescents in Niger household visit intervention on modern contraceptive use via perceived inequitable gender norms among husbands of married adolescent girls in Dosso, Niger at 24-month follow-up (2017-2019) ^a^**Boyce SC**, Deardorff J, Minnis AM, McCoy S, Shakya H, Challa S, Brooks M, Alion S, NouhouA, Silverman JG. Effect of a small group gender-synchronized family planning intervention on perceived gender norms among husbands of married adolescent girls in a cluster randomized control trial in Dosso, Niger. *Under review*.

### Mediation effects

The mediation pathway through gender inequitable social norms appears to have contributed to the observed association between the small group RMA intervention and reductions in IPV prevalence (Table 2 and Figure 2). Specifically, the estimated natural direct effect of the RMA intervention on IPV is a 6% reduction in IPV (95% CI: -0.20, -0.02). The estimated natural indirect effect of the RMA intervention via inequitable gender norms is a 2% reduction (95% CI: -0.07, 0.12) in IPV. This indirect effect suggests that 22.3% of the total effect between the RMA small group intervention and reduced prevalence of IPV is explained by a reduction in inequitable gender norms.

In both household and combination arms of the study, the natural direct pathways appear to account for the positive total effects on contraceptive use. For the household visits, the estimated natural direct effects is a 32% increase (95% CI: 0.13, 0.44) in current contraceptive use, 60% higher than the total effect. The estimated natural indirect effect via gender inequitable norms is an 11% decrease in contraceptive use (95% CI: -0.18, -0.01), suggesting that this pathway may have reduced the total effects of this intervention on contraceptive use (Table 2 and Figure 3). Similarly in the combination intervention, the estimated natural direct effect is a 28% increase (95% CI: 0.12, 0.46), 47% higher than the total effect for this arm, and the estimated natural indirect effect via gender inequitable norms is a 9% decrease (95% CI: -0.20, 0.02) in contraceptive use (Table 2).

## Discussion

This study examined the mechanistic role of inequitable gender norms on the observed total effects of the RMA intervention components on reductions in IPV and increases in current modern contraceptive use using data from a cRCT in Niger. Silverman et al. previously reported the estimated main effects of each RMA component on these outcomes, and Boyce et al. on the estimated effects of RMA components on inequitable gender norms (28, 29). This study extends this line of inquiry by providing evidence that reductions in perceived inequitable gender norms in the small groups RMA intervention may account for about one-quarter (22.3%) of the 8% reductions in IPV prevalence associated with assignment to this intervention component. Such results suggest that husbands’ perceptions of inequitable gender norms are an important part of the causal pathway between the small group intervention and behavior change around IPV perpetration among husbands. Findings for modern contraceptive use also underscore the importance of husbands’ perceptions of inequitable gender norms. Previous results suggest that the household intervention increased perceived *in*equitable gender norms among husbands and increased modern contraceptive use. The present mediation results demonstrated that the increases in modern contraceptive use would have been larger had the household intervention not increased inequitable gender norms. They also emphasize that the increases in modern contraceptive use in both the household and combination arm were achieved through other mechanisms than through reducing inequitable gender norms.

To our knowledge, this study is one of the first to provide rigorous evidence from a cluster randomized control trial of an intervention suggesting social norms change was one of the mechanisms behind reductions in intimate partner violence. *SASA!*, a social norms-shifting intervention that reduced IPV and HIV in Uganda, is one of very few interventions with evidence from a cRCT of both improving gender norms (measured by aggregating individual attitudes at the community level, a proxy of social norms) and health outcomes, suggesting that health outcomes changed via shifts in social norms, notwithstanding limitations of the social norms measurement used (25, 41). Similarly, *Program H* implemented in India demonstrated positive shifts in gender equity and IPV perpetration in a quasi-experimental trial (42). Neither of these studies, however, assessed for mediating effects of these social norms. In contrast, the *Tostan* model in Senegal both improved social norms and reduced female genital cutting (FGC), and while quasi-experimental evidence suggests messaging diffused through social networks, the final evaluation did not support the hypothesis that changes in social norms drove reductions in FGC (24). The present study, in consideration of our previously published results, contributes experimental evidence that the small group RMA intervention reduced IPV partially via decreasing perceived inequitable gender norms (28, 29). Not only do these results open the “black box” around how the RMA small group intervention may create behavior change to help inform its future use, they provide evidence supporting behavior change theories and frameworks that postulate the importance of changing underlying social norms in order to reduce IPV (8, 11-14).

For household visits and the combination arms, increases in inequitable gender norms associated with the household visits had a diminishing effect on current modern contraceptive use. These findings also affirm the importance of husbands’ perceived gender norms in shaping contraceptive use in this population; as inequitable gender norms increased, contraceptive use decreased. These changes in norms appear to have diminished what would have otherwise been larger increases in contraceptive use associated with receipt of the household and combination interventions; the estimated natural direct effects were stronger than the total effects for the household and small group arm.

These findings also raise important questions about the other mechanisms through which the RMA intervention components decreased IPV and increased contraceptive use. Because changes in norms did not contribute to the observed increase in contraceptive use and only partially explain decreases in IPV, other mechanisms must contribute to the interventions’ mechanisms. Other potential mechanisms for contraceptive use that warrant further study include social norms more proximal to acceptability of modern contraceptive use, husband and wife attitude change around acceptability of contraceptive use, husband and wife communication around contraceptive use, improved knowledge regarding contraceptive use, or improved access to products, all of which were goals of the intervention. Using data from this cRCT, Challa et al. found that assignment to the RMA intervention was associated with increases in spousal communication about contraceptive use and offers preliminary evidence that spousal communication may be a mechanism behind increases in contraceptive use (43). The mediation analysis combined all intervention arms into one group, so further assessment is needed to explore how this mechanism functions for each intervention arm, as each arm’s association with contraception use varied (e.g., no relationship with small groups) and this study suggests mediators also vary drastically by arm. Other potential mechanisms for IPV to consider include social norms more proximal to the social acceptability of IPV, individual attitudes regarding justification of IPV, and family and peer IPV behavior, all of which were targeted by the RMA intervention. Additional mediation analyses exploring these factors would be useful to furthering understanding of how the RMA components are impacting IPV and contraceptive use behavior.

The main (i.e., total) effects reported by Silverman et al. for the RMA intervention components indicate that the small groups (Arm 2) had statistically significantly reduction on the relative risk for IPV and that the household visits (Arm 1) and the combination (Arm 3) had statistically significant effects on contraceptive use. These associations were estimated as relative risks and are larger effects than observed in this study, which estimated effects on the linear scale to provide prevalence differences. These estimates of absolute measures of association provide a more conservative estimate of these relationships, and yet, the associations still remain (though diminished), further demonstrating the effects of the RMA intervention components on these outcomes.

These results should be considered in light of limitations. First, measurement of social norms is an emerging field and is rarely done in gender equity-related research. As such, the scale used in this study is new and has not been widely tested. It does, however, have modest reliability (alpha=0.62) and is one of the only scales available that measures gender norms, rather than a proxy based on individual attitudes, and therefore contributes to this nascent area of scientific research (10). Secondly, there is a lack of precision in the total effect and natural direct effect estimates and limited power to detect natural direct and indirect effects, particularly for IPV, despite the moderate sample size for this cRCT. The inverse odds ratio weighting approach using bootstrapping is known to provide less precise estimates than parametric approaches to mediation, yet this approach was chosen because it could accommodate the difference-in-differences regression model without violating assumptions (39). Finally, this mediation analyses assumes no uncontrolled confounding between 1) the exposure and outcome, 2) the exposure and mediator, and 3) the mediator and outcome. Verifying and upholding these assumptions is difficult in most mediation analysis, including this one. While the exposure assignment is theoretically ignorable given it was randomized, baseline imbalances across study arms were detected. While models controlled for these baseline imbalances that were also hypothesized to relate to the mediator and outcomes, it is possible there remains some uncontrolled confounding of the exposure-outcome and exposure-mediator relationship, especially given it was not feasible to measure the mediator more temporarily proximal to the exposure (39).

## Conclusion

This study is among the first cluster RCTs implemented in a LMIC that provides evidence suggesting that changes in inequitable gender norms were on the causal pathway to reductions in IPV. Furthermore, this study offers critical evidence to support theoretical hypotheses that reductions in inequitable gender norms are a potential mechanism behind interventions that reduce risk for IPV. In particular, these results provide a window into one way that the RMA intervention may have achieved reductions in IPV in the small group arm by reducing perceived inequitable gender norms among husbands of adolescent wives. The results from this study may help inform further scale up of the RMA small group model in Niger or contexts with similar social norms, and offer potential opportunities for further reducing IPV by enhancing the social norm-changing components of the intervention.

Additionally, these findings offer evidence to support theoretical hypotheses that higher levels of inequitable gender norms reduce the likelihood of modern contraceptive use. Further mechanistic research is needed to understand other mechanisms that contributed to reductions in IPV in the small groups and how the household visits and combination arms increased modern contraceptive use among married adolescent girls and their husbands, despite the negative effect of increases in perceived inequitable gender norms. Continued research is needed to understand how men can be engaged in efforts to improve upstream inequitable gender norms in a way that leads to improvements in IPV and modern contraceptive use, as well as a wide range of other downstream gender equity-related health outcomes.

## Data Availability

All data produced in the present study are available upon reasonable request to the authors

## Acknowledgements

We would like to thank the residents of the Dosso, Doutchi, and Loga districts of Niger who made themselves available to participate in this study and the Niger Ministry for Health in their facilitation of this research. We would also like to thank our local community partners at Pathfinder International and OASIS who were central to the implementation of this research. The first author’s work on this manuscript was supported by Eunice Kennedy Shriver National Institute of Child Health and Human Development of the National Institutes of Health (grant F31HD100019-01; PI: SC Boyce). The content is solely the responsibility of the authors and does not necessarily represent the official views of the National Institutes of Health. The research study from which data were utilized was funded by the Bill and Melinda Gates Foundation (OPP1195210, Prime: Pathfinder International; Research PI: J Silverman).

